# A whole population-based cohort study of the trajectory of the prevalence and the incidence of mental illness, challenging behaviour, and psychotropic medication prescribing in adults with intellectual disabilities in the Czech Republic between 2010 and 2022

**DOI:** 10.1101/2025.03.09.25323607

**Authors:** S Deb, J Jarkovský, H Melicharová, D Holub, B Limbu, P Třešňák

## Abstract

**Background:** It is essential to understand using a sizeable, centralised health database, the complex relationship among mental illness, challenging behaviour and psychotropic medication prescribing to help reduce the inappropriate overmedication of adults with intellectual disabilities. Diagnosing mental illness in adults with intellectual disabilities could be difficult because of the communication challenges and sometimes atypical manifestation, for example, through challenging behaviour.

**Method:** We explored the change in prevalence between 2010 and 2022 and incidence of mental illness, challenging behaviour and psychotropic medication prescribing between 2015 and 2022 among adults with intellectual disabilities and the relationships among them using a national health register in the Czech Republic. We collected data from the Institute of Health Information and Statistics of the Czech Republic.

**Results:** 62,636 (54% males) adults with intellectual disabilities contributed 704,503 person-years of data. In 2010, the prevalence of overall mental illness was 15·7%, challenging behaviour 29% and any psychotropic prescription 55%, increasing significantly by 2022 to 17·3%, 30·5% and 59%, respectively. The prevalence of overall mental illness, anxiety disorder, autism, ADHD and challenging behaviour increased significantly between 2010 and 2022. On the other hand, the incidence of newly diagnosed cases of most mental illnesses and challenging behaviour decreased significantly between 2015 and 2022. In contrast, the incidence of autism, ADHD, sleep disorders, and dementia increased significantly between 2015 and 2022. The incidence of challenging behaviour correlated significantly with psychoses, bipolar, anxiety and personality disorders. The prevalence of most psychotropic prescriptions increased significantly from 35% for antipsychotics, 17% for antidepressants, and 23% for mood stabilisers in 2010 to 37% for antipsychotics, 25% for antidepressants, and 28% for mood stabilisers in 2022 apart from anxiolytics which decreased from 14% in 2010 to 13% in 2022, despite a drop in the incidence of new prescriptions. The rate of psychotropic prescribing correlated significantly with the rate of mental illness and challenging behaviour. Among those receiving antipsychotics, only 18% in 2010 and 19% in 2022 had severe mental illness for which antipsychotics are indicated. Therefore, in >80% of cases, antipsychotic medications were prescribed off-label primarily for challenging behaviour. Similar trends were observed for other psychotropics. For example, mood stabilisers and antidepressants were prescribed more often for challenging behaviour in 38%-40% and 33%-40% of cases, respectively, than bipolar disorder (1%) and depression (8%-11%), respectively. In 2010, among those with challenging behaviour, 82% were prescribed any psychotropic medications, 62% antipsychotics, 20% antidepressants, and 30% mood stabilisers, increasing significantly by 2022 to 87%, 63%, 33%, and 37%, respectively. However, the prevalence of anxiolytic prescribing decreased significantly from 17% in 2010 to 14% in 2022. The incidence rates of new prescriptions for challenging behaviour fell by 1·1% for any psychotropic medications, 0·8% for antipsychotics, 0·7% for antidepressants and 0·1% for anxiolytics, but increased by 0·3% for hypnotics/sedatives and 0·4% for mood stabilisers.

**Conclusions:** The prevalence of mental illness and challenging behaviour increased over 12 years despite a drop in their incidences. This may indicate the chronic nature of these conditions. Challenging behaviour was significantly associated with some mental illnesses. Both mental illness and challenging behaviour were significantly associated with psychotropic medication prescribing. The prevalence of all psychotropic medication prescribing overall and especially for challenging behaviour was high and increased significantly over 12 years. This probably indicates long-term use of these medications. Antipsychotic medications were prescribed for severe mental illness only in a minority of cases. This indicates an off-licence use of antipsychotic medications in a very high proportion of cases. This trend of off-licence use was also evident in other psychotropic medications than antipsychotics. On the contrary, these medications were primarily used for challenging behaviour. The incidence of new prescriptions of antipsychotics, antidepressants, and anxiolytics decreased over the years, possibly at the expense of increased new prescriptions of mood stabilisers, primarily antiepileptics and also hypnotics/sedatives. The reasons for the significant increase in already high psychotropic medication prescription over 12 years and the trend of increase in new prescriptions of mood stabilisers and hypnotics/sedatives require scrutiny.

## Introduction

About 0·5-0·7% of the general adult population have intellectual (learning) disabilities (Carey et al., 2016; NHS Digital, 2021; Maulik et al., 2022). Intellectual disabilities are a group of etiologically diverse conditions originating during the developmental period, characterised by significantly below-average intellectual functioning and adaptive behaviour (WHO, 2004). Like the general population, adults with intellectual disabilities are prone to a whole range of mental illnesses. However, diagnosing mental illness in adults with intellectual disabilities could be difficult because of the communication challenges and sometimes atypical manifestation, for example, through challenging behaviour (Deb et al., 2022a; Deb et al., 2022b). It is, therefore, essential to understand the relationship between mental illness and challenging behaviour in this population using a sizeable, centralised health database. Otherwise, there is a danger of inappropriate overuse of psychotropic prescriptions for challenging behaviour. The overmedication of adults with intellectual disabilities is a significant public health concern (de Kuijper et al., 2010; Sheehan et al., 2015; Mehta & Glover, 2019; Song et al., 2023) as the RCT-based evidence for the efficacy of psychotropics for challenging behaviour in the absence of a mental illness among adults with intellectual disabilities is weak (see Deb, 2024 for a review), but the evidence of harm caused by the medication’s adverse effects is substantial (Ramerman et al., 2018; Deb et al., 2023; Sun et al., 2023)). The World Psychiatric Association (WPA) (Deb et al., 2009) and the UK NICE (NICE, 2015) have developed guidelines to address this issue along with the NHS England initiative STOMP (STopping Over-Medication of People with ID, autism or both) (Branford et al., 2018). All these guidelines recommend the use of non-pharmacological interventions such as positive behaviour support (PBS) (Gore et al., 2022) first before prescribing psychotropic medications for challenging behaviour. However, there has not been a longitudinal study to assess the impact of these guidelines on psychotropic prescribing for mental illness and challenging behaviour.

Therefore, we studied the change in the prevalence and incidence of mental illness, challenging behaviour, and psychotropic medication prescribing using the International Classification of Diseases-10^th^ revision (ICD-10) (WHO, 2004) in adults with intellectual disabilities between 2010 and 2022 in the Czech Republic to assess the impact of NICE and WPA guidelines and the UK STOMP initiative on psychotropic medication prescribing in this population and its relationship with mental illness and challenging behaviour.

## Method

We collected data from the National Registry of Reimbursed Health Services (NRRHS) in the Czech Republic. NRRHS contains all healthcare data provided under public health insurance, which covers almost 100% of the population in the Czech Republic. NRRHS is part of the National Health Information System (NHIS) regulated by the Institute of Health Information and Statistics of the Czech Republic (“IHIS CR“ or “the Institute“). The Ministry of Health in the Czech Republic established the Institute in 1960 to implement the Health Services Act. The Institute follows the principles of the European Statistics Code of Practice, which represents a common summary of European standards designated for statistical organisations and the whole European statistical system to secure a high quality and credibility of European data. In the Czech Republic, all adults are registered with this national registry to receive reimbursement for their health service costs. The health data such as the diagnosis, treatment and diagnostic tests on all adults in the country who have come across any health service such as in-patient, outpatient or community provided by any specialities such as medicine, surgery, psychiatry, including health professionals like doctors including general practitioners and nurses and other allied health professionals like physiotherapists, psychologists, occupational therapists, radiologists, haematologists, etc. The International Classification of Diseases-10^th^ revision (ICD-10) (WHO, 2004) criteria are used by all who input data into the national register. The prevalence data were collected from 2010, the year after the WPA international guideline (Deb et al., 2009) was published, until 2022, the last year ICD-10 diagnoses were made, as ICD-11 (WHO, 2019) was published in 2022. The incidence rates of the first occurrence of episodes within the last five years were collected from 2015 to 2022, allowing for the recording of only new entries that were not present prior to 2015.

We have identified adults with intellectual disabilities using the ICD-10 codes of F70-79 from any service, either as a primary or a secondary diagnosis. The intellectual disabilities diagnosis was subdivided into F70 mild intellectual disabilities, F71 moderate intellectual disabilities, F72 severe intellectual disabilities, F73 profound intellectual disabilities, F78 other intellectual disabilities, and F79 unspecified intellectual disabilities. The diagnosis of challenging behaviour was made using the ICD-10 codes F70x.1 (significant impairment of behaviour requiring attention or treatment) and F70x.8 (other impairments of behaviour). A psychiatric diagnosis is recorded using the following ICD-10 codes. F20-29: schizophrenia, schizotypal and delusional disorders (psychoses); F30-31: bipolar disorder; F32-39: depressive disorder; F40-48: anxiety disorders; F51: sleep disorders; F60-69: personality disorders; and F00-03: dementia. We used F84 for autism spectrum disorder and F90 for ADHD diagnosis. To compare our findings with the previous English general practice register-based study (Sheehan et al., 2015), we recorded a diagnosis of a severe mental illness if someone had a diagnosis of F20-29 (psychosis) and/or F30-31 (bipolar disorder) and also a diagnosis of any psychiatric disorder if someone has any of the following ICD-10 diagnosis; F20-48 (psychoses, bipolar disorder, depressive disorder and anxiety disorders).

We have utilised the WHO Anatomical Therapeutic Chemical (ATC) codes and the Therapeutic Chemical Classification systems (www.whocc.no/atc-do-index, accessed on 25.03.2025) to classify psychotropic medication groups. The following codes were used. N05A: antipsychotics; NO6A: antidepressants; NO5B: anxiolytics (anti-anxiety medications); NO5C: hypnotics/sedatives; NO3: Anti-epileptic medication; NO5AN: lithium. Anyone receiving NO5AN, NO3AX, NO3AF or NO3AG is recorded as receiving a mood stabiliser. Anyone receiving any of these medications was recorded as receiving any psychotropic.

## Statistical analysis

The data were processed using the Vertica 9 database and analysed using SPSS 29.0.1.0 (IBM Corporation 2023) and STATA/IC 15.1.

Absolute and relative frequencies were used as standard descriptive statistics for categorical data. The incidence rate ratio (IRR) and its statistical significance were adopted to compare the occurrence of psychotropic prescriptions between years. Binary logistic regression analyses were used to determine factors influencing the occurrence of psychotropic medication, mental illness, and challenging behaviour among adults with intellectual disabilities. We estimated models using the stepwise forward conditional method. If adults with intellectual disabilities received a new prescription of psychotropic medication or a diagnosis of a mental illness or challenging behaviour after cohort entry (that is, during follow-up), we considered them no longer at risk for that psychotropic prescription or mental illness or challenging behaviour and removed them from the cohort.

We conducted multivariate regression analysis using the incidences of mental illnesses as dependent variables and age, gender, severity of intellectual disabilities, and incidence of challenging behaviour as the independent variables. We also conducted multivariate regression analysis using the incidences of new prescriptions of different psychotropic medications as dependent variables and gender, age, severity of intellectual disabilities, and incidences of challenging behaviour and different mental illnesses as the independent variables. We used Wald tests to assess the overall significance of categorical variables and categorical interaction terms. We considered a p-value of 0.05 to be statistically significant (two-tailed).

## Patient and public involvement

Children of the Full Moon, a non-governmental parent organisation for people with autism spectrum disorder and intellectual disabilities in the Czech Republic, has been involved at every stage of the study, from conceptualisation through design, data analysis, and paper writing. The organisation’s chairman (PT), also the parent of a child with autism, is a co-author.

## Results

Table 1 shows the prevalence (2010 and 2022) and the incidence rates of new cases (2015 and 2022) of different mental illnesses, challenging behaviours and neurodevelopmental disorders like autism and ADHD. In 2010, 53,551 adults had a diagnosis of intellectual disabilities. This figure increased to 58,678 in 2015 and 62,636 in 2022. The incidence rate is presented per 10,000 person-years to compare data with the previous English study (Sheehan et al., 2015). Figure 1 illustrates the trajectory of new cases recorded for various mental illnesses and challenging behaviours from 2015 to 2022, showing a decline in the incidence of new cases over the years, except for a slight increase in new cases of any mental illness and anxiety disorder between 2021 and 2022.

**Figure 1.**
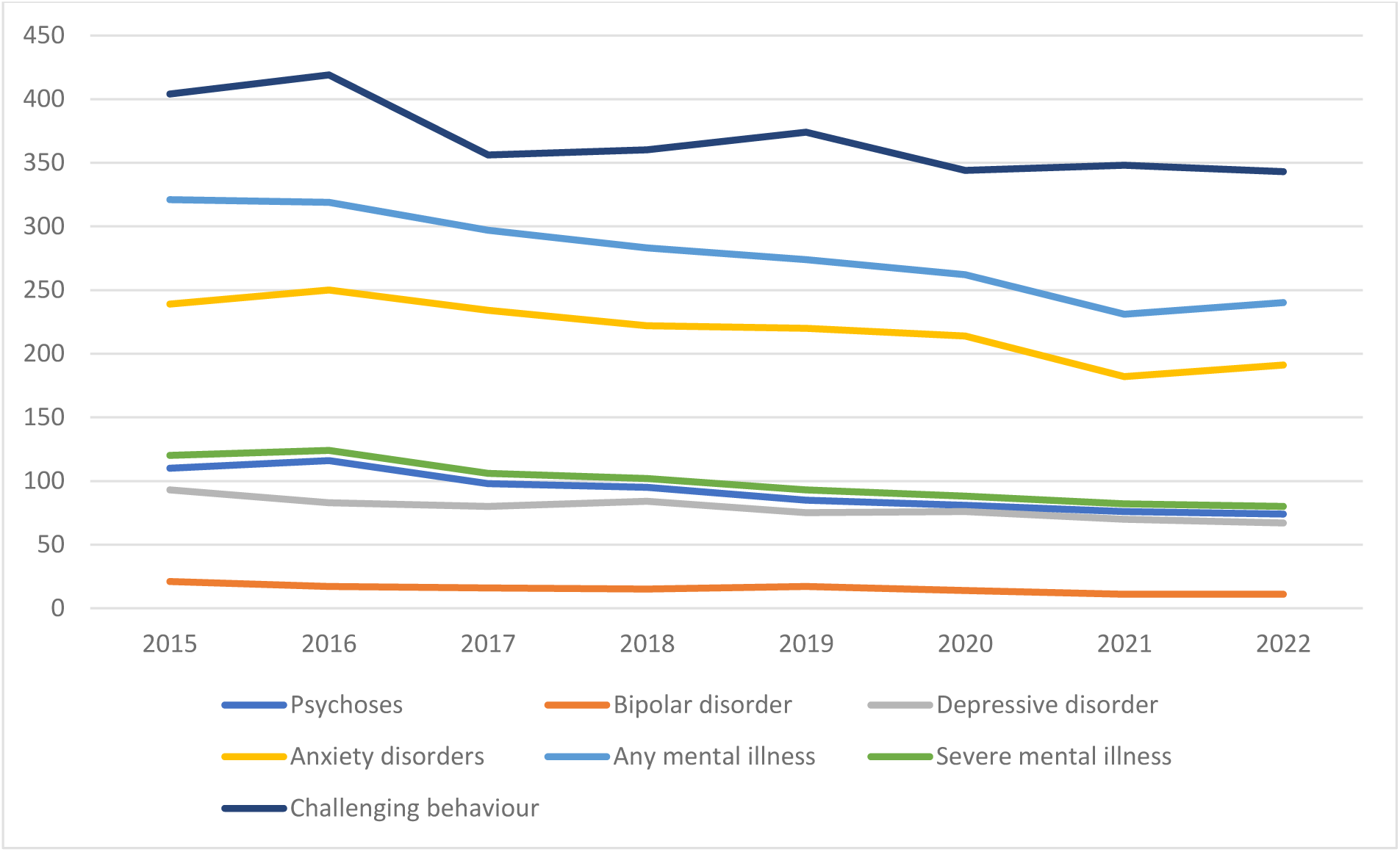
Incidence of mental illness and challenging behaviour per 10,000 person-years of adults with intellectual disabilities between 2015 and 2022

In 2010, the prevalence of overall mental illness was 15·7%, challenging behaviour 29% and any psychotropic prescription 55%, increasing significantly by 2022 to 17·3%, 30·5% and 59%, respectively. The prevalence of any mental illness and all individual mental illnesses, along with the neurodevelopmental disorders and challenging behaviour, increased between 2010 and 2022. The prevalence of any mental illness was 15.7%, anxiety disorder 7.4%, autism 0.7%, ADHD 0.5%, and challenging behaviour 29% in 2010, which increased significantly in 2022 to 17.3%, 9%, 3%, 2%, and 30.5%, respectively. On the other hand, the incidence of newly diagnosed cases of most mental illnesses and challenging behaviour decreased significantly between 2015 and 2022, apart from autism, ADHD, sleep disorder, and dementia, which increased significantly between 2015 and 2022.

**Table 1.** Prevalence (2010-2022) and incidence (2015-2022) of mental illness, challenging behaviour and neurodevelopmental disorders.

| **Diagnosis** | **Prevalence**<br>**(N=53,551)**<br>**2010 N (%)** | **Prevalence**<br>**(N=62,636)**<br>**2022 N (%)** | **Incidence**<br>**per 10,000**<br>**person-**<br>**years**<br>**(N=58,678)**<br>**2015** | **Incidence**<br>**per 10,000**<br>**person-**<br>**years**<br>**(N=62,636)**<br>**2022** | **Level of**<br>**significance**<br>**(p-value)** |
| --- | --- | --- | --- | --- | --- |
| Any mental illness | 8419<br>(15.7%) | 10819 (17.3%) | 321 | 240 | <0.001 |
| Severe mental illness | 3938 (7.4%) | 4835 (7.7%) | 120 | 80 | <0.001 |
| Psychoses | 3,773 (7%) | 4,560 (7.3%) | 110 | 74 | <0.001 |
| Bipolar disorder | 217 (0.4%) | 327 (0.5%) | 22 | 11 | <0.001 |
| Depressive disorder | 1,363 (3%) | 1,754 (3%) | 93 | 67 | <0.001 |
| Anxiety disorders | 3,956 (7.4%) | 5,300 (9%) | 239 | 191 | <0.001 |
| Dementia | 774 (2%) | 1548 (3%) | 73 | 86 | 0.01 |
| Personality disorder | 1,899 (4%) | 2659 (4.3%) | 102 | 77 | <0.001 |
| Sleep disorder | 102 (0.2%) | 169 (0.3%) | 9 | 11 | 0.359 |
| Autism | 370 (0.7%) | 1854 (3%) | 17 | 26 | <0.001 |
| ADHD | 244 (0.5%) | 937 (2%) | 11 | 18 | 0.002 |
| Challenging behaviour | 15,512<br>(29%) | 19,094<br>(30.5%) | 404 | 343 | <0.001 |

Table 2 shows the prevalence (2010 and 2022) and incidence (2015 and 2022) data on the overall prescription of psychotropic medications and the rate of psychotropic medications prescribed for challenging behaviour. We also presented prevalence data (2010 and 2022) on the proportion of those who received different psychotropic medications with different mental illnesses or challenging behaviours.

The prevalence of psychotropic medication prescribing among adults with intellectual disabilities increased significantly from 55% in 2010 to 59% in 2022. Similarly, antipsychotic prescribing increased significantly from 35% in 2010 to 37% in 2022, and antidepressants from 17% in 2010 to 25% in 2022. The prevalence of mood stabiliser prescriptions increased significantly from 23% in 20210 to 28% in 2022, and hypnotics/sedatives from 0.5% in 2010 to 1% in 2022. In contrast, the prevalence of anxiolytic prescribing fell significantly from 14% in 2010 to 13% in 2022.

Among those who showed challenging behaviour, a high proportion received psychotropic medications (82%) in 2010, significantly increasing in 2022 to 87%. A high proportion also received antipsychotic medications (62%) in 2010, which increased significantly in 2022 to 63%. Similarly, the prevalence of antidepressant prescriptions for challenging behaviour increased significantly from 20% in 2010 to 33% in 2022, and mood stabilisers from 30% in 2010 to 37% in 2022. In contrast, the prevalence of anxiolytic prescriptions for challenging behaviour decreased significantly from 17% in 2010 to 14% in 2022.

**Table 2.** Prevalence (2010-2022) and incidence (2015-2022) of psychotropic prescribing.

| **The overall rate of psychotropic prescription** |  |  |  |  |  |
| --- | --- | --- | --- | --- | --- |
| Psychotropics | Prevalence<br>(N=53,551)<br>2010 N (%) | Prevalence<br>(N=62,636)<br>2022 N (%) | Incidence per<br>10,000 person-years (N=58,678)<br>2015 | Incidence<br>per 10,000<br>person-years<br>(N=62,636)<br>2022 | Level of<br>significance<br>(p-value) |
| Any psychotropic | 29,616<br>(55%) | 36,981 (59%) | 457 | 383 | <0.001 |
| Antipsychotics | 18,808<br>(35%) | 23,001 (37%) | 317 | 257 | 0.469 |
| Antidepressants | 9,397 (17%) | 15,847 (25%) | 334 | 278 | 0.146 |
| Anxiolytics | 7,705 (14%) | 7,953 (13%) | 326 | 297 | 0.05 |
| Hypnotics/sedatives | 290 (0.5%) | 635 (1%) | 51 | 71 | <0.001 |
| Mood stabilisers | 12,158<br>(23%) | 17,488 (28%) | 379 | 368 | <0.001 |
| **The rate of psychotropics prescribed for challenging behaviour** |  |  |  |  |  |
| Psychotropics | Prevalence<br>(N=15,512)<br>2010 N (%) | Prevalence<br>(N=19,094) 2022<br>N (%) | Incidence per<br>10,000 person-years (N=17,015)<br>2015 | Incidence<br>per 10,000<br>person-years<br>(N=19,094)<br>2022 | Level of<br>significance<br>(p-value) |
| Any psychotropic | 12,689<br>(82%) | 16,601 (87%) | 507 | 403 | 0.004 |
| Antipsychotics | 9,590 (62%) | 12,078 (63%) | 512 | 427 | 0.026 |
| Antidepressants | 3,098 (20%) | 6,378 (33%) | 476 | 413 | 0.115 |
| Anxiolytics | 2,693 (17%) | 2,658 (14%) | 353 | 338 | 0.02 |
| Hypnotics/sedatives | 68 (0.4%) | 213 (1%) | 45 | 74 | 0.01 |
| Mood stabilisers | 4,634 (30%) | 6,987 (37%) | 370 | 414 | <0.001 |
| **The rate of mental illnesses and challenging behaviour among those who received different psychotropic medications** |  |  |  |  |  |
| **Mental illness** |  |  | **Challenging behaviour** |  |  |
| Psychotropics | 2010 | 2022 | Psychotropics | 2010 | 2022 |
| Antipsychotics for severe mental illness | 3475 (18%) | 4440 (19%) | Antipsychotics | 9590 (51%) | 12078 (52%) |
| Antidepressants for depressive disorder | 996 (11%) | 1300 (8%) | Antidepressants | 3098 (33%) | 6378 (40%) |
| Anxiolytics for anxiety disorders | 1232 (16%) | 1481 (19%) | Anxiolytics | 2693 (35%) | 2658 (33%) |
| Hypnotics/sedatives for sleep disorders | 0 | 2 | Hypnotics/sedatives | 68 (23%) | 212 (33%) |
| Mood stabilisers for bipolar disorder | 137 (1%) | 204 (1%) | Mood stabilisers | 4634 (38%) | 6987 (40%) |

The incidence rates of new cases of psychotropic prescriptions are presented in Table 2 and Figure 2. The incidence rates decreased from 2015 to 2022 for most psychotropic medication prescriptions, apart from hypnotics/sedatives, which increased between 2015 and 2022. The trajectory showed a rise in incidence in most psychotropics in 2016, followed by a subsequent decrease in 2017. However, there was another rise in the incidence rates of most psychotropic medications in 2022 (the last year of data collection) (see Figure 2).

**Figure 2.**
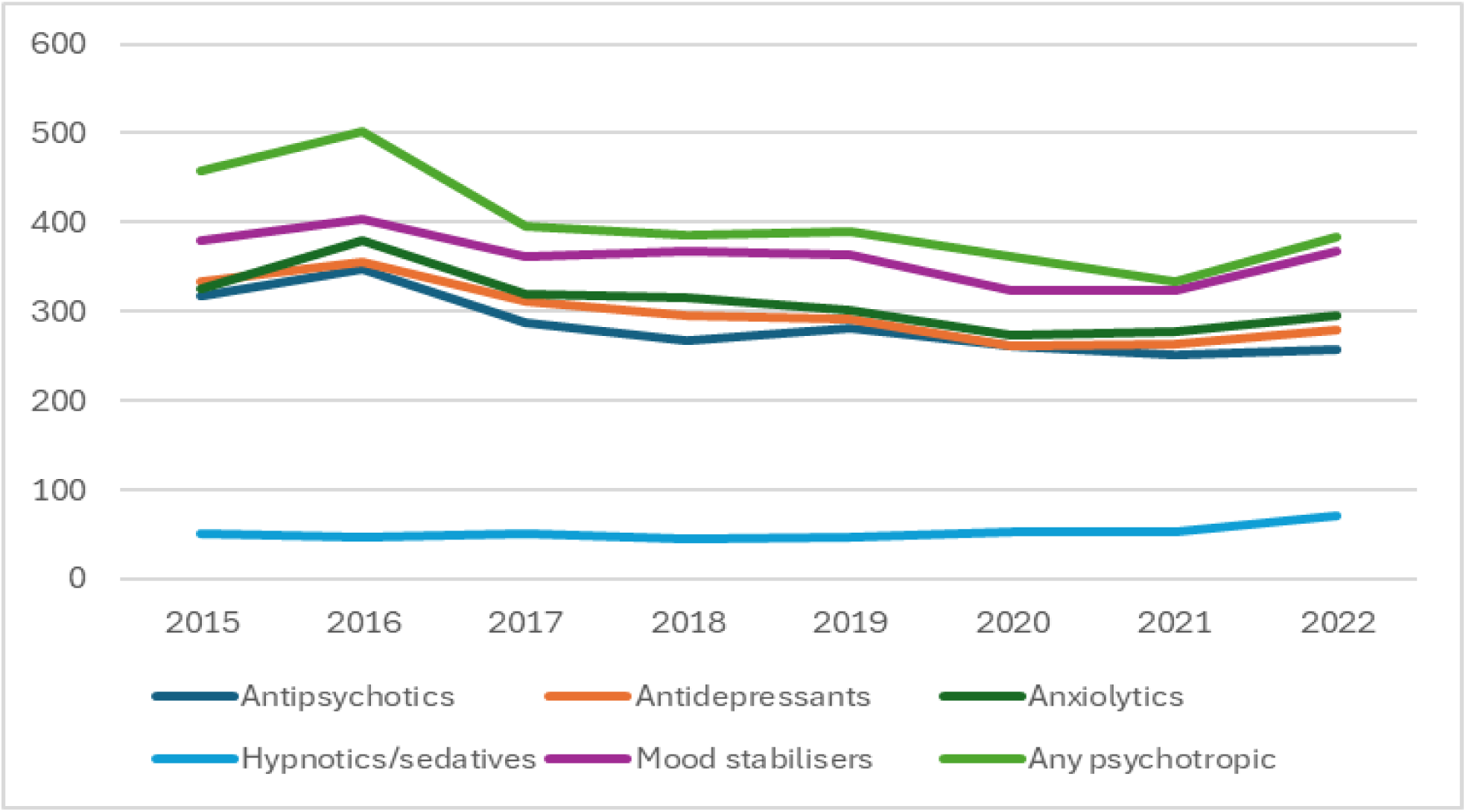
Incidence of new psychotropic prescriptions between 2015 and 2022 per 10,000 person-years of adults with intellectual disabilities

Whereas the incidence rate of new prescriptions of antipsychotics, antidepressants and anxiolytics decreased between 2015 and 2022 for those who displayed challenging behaviour, the rate of new prescriptions of hypnotics/sedatives and mood stabilisers increased significantly over the same period (see Table 2). Figure 3 shows that the incidence rates of new prescriptions for psychotropic medications to address challenging behaviour decreased during 2017-2018 and again in 2020-2021, with corresponding increases in 2019-2020 and 2022.

**Figure 3.**
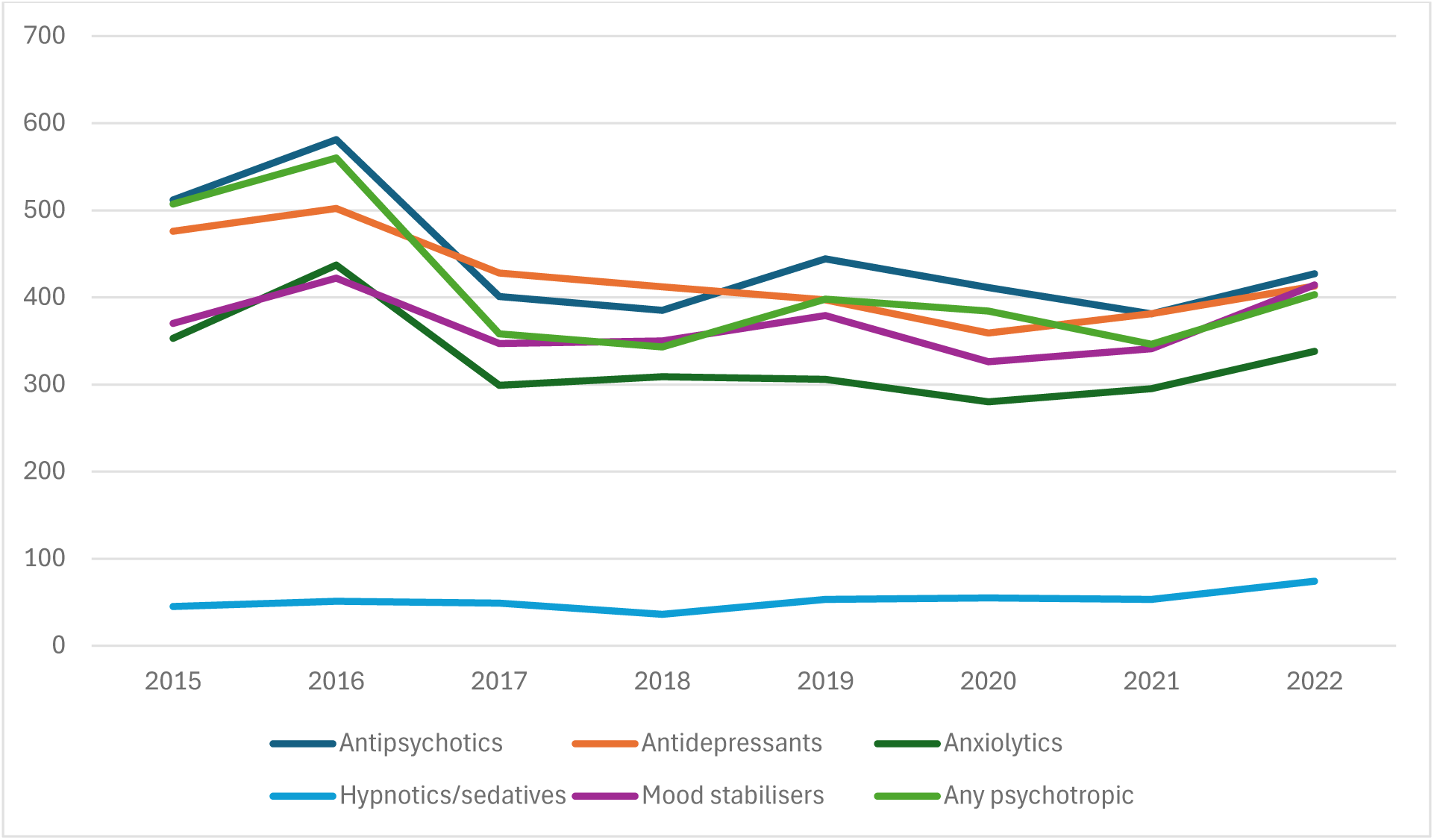
Incidence of new prescriptions of psychotropic medications for challenging behaviour in adults with intellectual disabilities between 2015 and 2022 per 10,000 person-years

Among those who received antipsychotics, only 18% in 2010 and 19% in 2022 had a diagnosis of severe mental illness for which they are indicated (see Table 2). This means that in over 80% of cases, antipsychotics were prescribed when there was no severe mental illness present. On the contrary, among those who received antipsychotics, 51% in 2010 and 52% in 2022 had a diagnosis of challenging behaviour. Antidepressants were prescribed for depressive disorders only for 11% in 2010 and 8% of cases in 2022. On the other hand, antidepressants were prescribed in 33% of cases in 2010 and 40% in 2022 for challenging behaviour. Similarly, anxiolytics were used for anxiety disorders only for 16% in 2010 and 19% of cases in 2022 but for challenging behaviour in 35% of cases in 2010 and 33% in 2022. Similarly, mood stabilisers were prescribed for challenging behaviour in 38% of cases in 2010 and 40% in 2022. In contrast, mood stabilisers were prescribed for only 1% of cases of bipolar disorder in 2010 and 2022 (see Table 2).

Table 3 shows the demographic characteristics of all adults with a diagnosis of intellectual disabilities in 2022 (n=62,636), new cases of adults with intellectual disabilities who showed challenging behaviour (n=2147), and those receiving new prescriptions of any psychotropic medications in 2022 (n=2398). Demographic distribution showed a higher rate of males, a lower age group (<40 years) and mild intellectual disabilities among those who have intellectual disabilities, but also those who showed challenging behaviour and received any psychotropic medications.

**Table 3.** Characteristics of the study population and psychotropic medicine prescription.

| **All adults with intellectual disabilities in 2022 (n=62,636) (F70-74)** |  |
| --- | --- |
| Gender |  |
| Male | 34 014 (54%) |
| Female | 28 622 (46%) |
| Age group |  |
| <40 years | 29 226 (47%) |
| 40-64 years | 24 453 (39%) |
| ≥ 65 years | 8 957 (14%) |
| The severity of intellectual disabilities |  |
| Mild and moderate | 49 032 (78%) |
| Severe and profound | 7 998 (13%) |
| Other and unspecified | 5 606 (9%) |
| **Incidence of those who displayed challenging behaviour in 2022 (F70x.1 and/or x.8) (n=2147)** |  |
| Gender |  |
| Male | 1 137 (53%) |
| Female | 1 010 (47%) |
| Age group |  |
| <40 years | 915 (42.6%) |
| 40-64 years | 843 (39.3%) |
| ≥ 65 years | 389 (18.1%) |
| The severity of intellectual disabilities |  |
| Mild and moderate | 1 791 (83.4%) |
| Severe and profound | 307 (14.3%) |
| Other and unspecified | 49 (2.3%) |
| **Incidence of any new psychotropic medication prescription in 2022 (n=2,398)** |  |
| Gender |  |
| Male | 1 320 (55%) |
| Female | 1 078 (45%) |
| Age group |  |
| <40 years | 1 115 (46.5%) |
| 40-64 years | 861 (35.9%) |
| ≥ 65 | 422 (17.6%) |
| The severity of intellectual disabilities |  |
| Mild and moderate | 1 851 (77.2%) |
| Severe and profound | 271 (11.3%) |
| Other and unspecified | 276 (11.5%) |

Table 4 shows the results of multivariate regression analysis. The incidence of challenging behaviour correlated significantly with the incidences of psychoses, bipolar, anxiety and personality disorders.

The female gender was significantly associated with the incidences of bipolar disorder (<0.005), depressive disorder (p<0.001), anxiety disorder (p<0.001), and personality disorder (p<0.001). Older age was significantly related to the incidences of psychoses (p<0.05), depressive disorder (p<0.001), anxiety disorder (p<0.001) and personality disorder (p<0.001). Severe and profound intellectual disabilities were significantly associated with the incidences of psychoses (p<0.001), anxiety disorder (p<0.005) and personality disorder (p<0.001).

**Table 4.** Factors influencing the occurrence of incidence of different mental illnesses among adults with intellectual disabilities (F70-F79) (multivariate analysis)

|  |  | Psychoses<br>(F20-29) |  | Bipolar disorder<br>(F30-31) |  | Depressive<br>disorder (F32-39) |  | Anxiety disorder<br>(F40-48) |  | Personality<br>Disorder (F60-69) |  |
| --- | --- | --- | --- | --- | --- | --- | --- | --- | --- | --- | --- |
|  |  | OR (95% CI) | p | OR (95% CI) | p | OR (95% CI) | p | OR (95% CI) | p | OR (95% CI) | p |
| Gender | Women | N/A | N/A | 1.259 (1.090;<br>1.454) | 0.002 | 1.507 (1.413;<br>1.608) | <0.001 | 1.337 (1.286;<br>1.390) | <0.001 | 0.820 (0.772;<br>0.871) | <0.001 |
| Age | < 40 | reference |  | reference |  | reference |  | reference |  | reference |  |
|  | 40–64 | 1.121 (1.015;<br>1.239) | 0.025 | N/A | N/A | 1.240 (1.114;<br>1.380) | <0.001 | 1.689 (1.581;<br>1.804) | <0.001 | 2.900 (2.546;<br>3.303) | <0.001 |
|  | ≥ 65 | 1.154 (1.044;<br>1.276) | 0.005 | N/A | N/A | 1.371 (1.233;<br>1.523) | <0.001 | 1.259 (1.177;<br>1.347) | <0.001 | 2.050 (1.795;<br>2.341) | <0.001 |
| The severity<br>of ID | Mild and<br>moderate | reference |  | reference |  | reference |  | reference |  | reference |  |
|  | Severe and<br>profound | 2.684 (2.243;<br>3.212) | <0.001 | 1.231 (0.913;<br>1.658) | 0.173 | 1.097 (0.980;<br>1.229) | 0.109 | 0.904 (0.845;<br>0.968) | 0.004 | 1.530 (1.331;<br>1.759) | <0.001 |
|  | Other and<br>unspecified | 1.172 (0.954;<br>1.442) | 0.131 | 0.889 (0.620;<br>1.276) | 0.524 | 0.565 (0.482;<br>0.661) | <0.001 | 0.391 (0.354;<br>0.432) | <0.001 | 0.417 (0.344;<br>0.506) | <0.001 |
| Challenging<br>behaviour |  | 2.529 (2.382;<br>2.686) | <0.001 | 2.085 (1.796;<br>2.420) | <0.001 | N/A | N/A | 1.129 (1.082;<br>1.179) | <0.001 | 1.626 (1.530;<br>1.729) | <0.001 |

Multivariate analysis of factors influencing the incidence of psychotropic medication and occurrence of incidence of psychiatric disorders among adults with intellectual disabilities with odds ratio and p-values. CI= Confidence Interval, N/A= Not applicable (no significant association was found), OR= Odds ratio, and P= p-values.

Table 5 presents the findings from a multivariate regression analysis, using the incidence of new psychotropic prescribing as the dependent variable. The rate of challenging behaviour was significantly associated with the rate of prescription of any psychotropic medication (p<0.001), antipsychotics (p<0.001), antidepressants (p<0.001), and anxiolytics (p<0.001). The female gender was statistically significantly associated with the prescription of antidepressants and anxiolytics but not antipsychotics and mood stabilisers. Older age (>65 years) was statistically significantly related to the prescription of all psychotropic medications. The more severe intellectual disabilities (severe and profound intellectual disabilities) were statistically significantly associated with the prescription of all psychotropics apart from antipsychotic medication. Anxiety disorder was statistically significantly associated with all psychotropic medication prescriptions.

**Table 5.** Factors influencing the incidence of psychotropic medication prescribing among adults with intellectual disabilities (F70-F79) (multivariate analysis)

|  |  | Antipsychotics |  | Antidepressants |  | Anxiolytics |  | Hypnotics and sedatives |  | Any psychotropic |  |
| --- | --- | --- | --- | --- | --- | --- | --- | --- | --- | --- | --- |
|  |  | OR (95% CI) | p | OR (95% CI) | p | OR (95% CI) | p | OR (95% CI) | p | OR (95% CI) | p |
| Gender | Women | N/A | N/A | 1.143 (1.105; 1.182) | <0.001 | 1.163 (1.126; 1.202) | <0.001 | N/A | N/A | N/A | N/A |
| Age | < 40 | reference |  | reference |  | reference |  | reference |  | reference |  |
|  | 40–64 | 0.729 (0.694; 0.767) | <0.001 | 0.920 (0.873; 0.970) | 0.002 | 1.092 (1.036; 1.152) | 0.001 | 0.545 (0.485; 0.612) | <0.001 | 1.035 (0.989; 1.083) | 0.138 |
|  | ≥ 65 | 0.688 (0.655; 0.724) | <0.001 | 0.825 (0.783; 0.870) | <0.001 | 1.098 (1.042; 1.158) | <0.001 | 0.788 (0.706; 0.880) | <0.001 | 0.937 (0.895; 0.980) | 0.005 |
| The severity of ID | Mild and moderate | reference |  | reference |  | reference |  | reference |  | reference |  |
|  | Severe and profound | N/A | N/A | 0.923 (0.868; 0.982) | 0.011 | 0.803 (0.760; 0.849) | <0.001 | 0.621 (0.552; 0.699) | <0.001 | 0.853 (0.812; 0.895) | <0.001 |
|  | Other and unspecified | N/A | N/A | 0.747 (0.690; 0.807) | <0.001 | 0.712 (0.663; 0.766) | <0.001 | 0.593 (0.507; 0.693) | <0.001 | 0.783 (0.735; 0.834) | <0.001 |
| Challenging behaviour |  | 2.198 (2.124; 2.276) | <0.001 | 1.892 (1.827; 1.960) | <0.001 | 1.101 (1.062; 1.142) | <0.001 | N/A | N/A | 1.167 (1.130; 1.205) | <0.001 |
| Psychoses |  | 1.357 (1.285; 1.434) | <0.001 | 1.184 (1.119; 1.254) | <0.001 | 1.107 (1.044; 1.173) | 0.001 | N/A | N/A | 0.728 (0.685; 0.774) | <0.001 |
| Bipolar disorder |  | 1.287 (1.087; 1.524) | 0.003 | N/A | N/A | N/A | N/A | N/A | N/A | 0.739 (0.600; 0.911) | 0.005 |
| Depressive disorder |  | 1.694 (1.572; 1.825) | <0.001 | 1.854 (1.729; 1.987) | <0.001 | 1.522 (1.412; 1.641) | <0.001 | 1.356 (1.121; 1.640) | 0.002 | N/A | N/A |
| Anxiety disorder |  | 2.602 (2.487; 2.722) | <0.001 | 3.259 (3.126; 3.398) | <0.001 | 2.216 (2.119; 2.316) | <0.001 | 2.035 (1.823; 2.272) | <0.001 | 1.606 (1.538; 1.678) | <0.001 |

Multivariate analysis of factors influencing the incidence of psychotropic medication and occurrence of incidence of psychiatric disorders among adults with intellectual disabilities with odds ratio and p-values. CI= Confidence Interval, N/A= Not applicable (no significant association was found), OR= Odds ratio, and P= p-values.

## Discussion

### Mental illness, neurodevelopmental disorders and challenging behaviour

Although the overall prevalence of mental illness increased significantly over the 12-year study period between 2010 (15·7%) and 2022 (17·3%), the prevalence of most individual mental illnesses remained similar, apart from anxiety disorder, which increased significantly from 7·4% to 9%. However, the prevalence of challenging behaviour, autism and ADHD have all also increased significantly over the 12 years. The prevalence of mental illnesses, neurodevelopmental disorders, and challenging behaviour among adults with intellectual disabilities varied widely in previous studies for obvious methodological reasons (Deb et al., 2022a). Ours is the first large national health register-based study that used ICD diagnosis, which had not been done previously for adults with intellectual disabilities using a large national health database. However, the overall prevalence of mental illness and challenging behaviour found in the current study is similar to what was reported in some previous studies (Deb et al., 2022a). For example, a whole population-based cohort study of adults with intellectual disabilities in Scotland found the overall prevalence (Henderson et al., 2020) of mental illness and severe mental illness among 16.3% and 6.2% of the cohort (n=1190), respectively, at study entry in 2002-2004 compared with 15.7% and 7.4% respectively in the current study at study entry in 2010 (n=53,551). Similarly, in the Scottish study, depressive disorder was diagnosed among 4.3%, anxiety disorder in 2.7%, autism in 7.6%, and ADHD in 1.3% of the cohort, compared with 3%, 9%, 3%, and 2%, respectively in the current study in 2022. The significant rise in the prevalence of anxiety disorders in the current study over the 12 years may be a reflection of clinicians’ better awareness over the last decade of the presence of anxiety disorders in adults with intellectual disabilities. Similarly, the significant rise in the prevalence of autism and ADHD during the same period is likely to be due to a better awareness of these diagnostic categories among clinicians.

The reported prevalence of challenging behaviour varied widely in previous studies because of the different methodologies used. A significant problem has been defining and diagnosing challenging behaviour in this population. The current study employed an ICD-10 diagnosis, which has not been used in previous studies, to diagnose challenging behavior, despite the fact that ICD-10 provides an explicit qualifier with an extended ‘x’ code for this diagnosis. Despite these methodological differences, the prevalence of challenging behaviour reported in the current study is similar to some earlier studies (Deb et al., 2022a, b). For example, in the Scottish population-based study (Henderson et al., 2020), 20.5% of the cohort at study entry exhibited challenging behavior, compared with 29% in the current study at study entry. The significant rise in the prevalence of challenging behaviour from 2010 to 2022 in the current study is difficult to explain. It may be the effect of the Covid-19 pandemic, which has shown increased challenging behaviour in most recent studies (Gil-Llario et al., 2023). There may be even greater awareness among clinicians of this diagnosis, or they may have used the ICD-10 codes more frequently for this diagnosis in 2022 than in 2010. However, the incidence rate of new challenging behaviour cases fell by 0·6% from 404 per 10,000 person-years in 2015 to 343 in 2022. An increased prevalence rate of challenging behaviour over the years, despite the decrease in the incidence rate, may be due to the chronic nature of challenging behaviour. The incidence rates of most conditions fell significantly apart from sleep disorders, autism and ADHD, all of which increased over seven years, and the increase in autism and ADHD rates was statistically significant.

## Psychotropic medication prescribing

### The prevalence of psychotropic medication prescribing among adults with intellectual disabilities between 2010 and 2022

The reported prevalence rates of psychotropic medication prescribing for adults with intellectual disabilities in previous studies varied from as low as 10% to as high as 67%, depending on the method of data collection (Song et al., 2023; Deb, 2024). In a recent meta-analysis of 24 studies, Song and colleagues (Song et al., 2023) reported a pooled prevalence of psychotropic prescribing in adults with intellectual disabilities. Pooled prevalence for any psychotropic prescription was 41% (95% confidence interval (CI) 35-46%), antipsychotics 31% (95% CI 27-35%), antidepressants 14% (95% CI 9-19%), anxiolytics 9% (95% CI 4-15%), and hypnotics/sedatives 5% (95% CI 2-8%) (Song et al., 2023). This shows that apart from the prevalence of overall psychotropic medications and hypnotics/sedatives prescription among adults with intellectual disabilities in the current study, the prevalence rates of other psychotropic medication prescriptions at the study entry were within the 95% confidence intervals reported in the recent meta-analysis (Song et al., 2023). In the English general practice register-based study of 33,016 adults with intellectual disabilities (Sheehan et al., 2015), the rate of any psychotropic prescription was 49% at study entry in 1999 and 63% at the study end in 2013. This compares with the 55% prevalence of any psychotropic prescription in 2010 and 59% in 2022 in the current study. The rate of increase in psychotropic medication prevalence in our study over 12 years was lower than in the English study over 15 years. The most commonly prescribed psychotropic medication was antipsychotics in 2022 (37%), followed by mood stabilisers, primarily antiepileptics (28%), antidepressants (25%), anxiolytics (13%) and hypnotics/sedatives (1%). This compares with a recent study among adults with intellectual disabilities in community homes and supported living accommodations in England and Wales in the UK which reported of 190 prescriptions 47% were antipsychotics followed by 23% antidepressants, 13% antiepileptics, 7% anxiolytics in the form of benzodiazepine (Deb et al., 2025).

The prevalence of antipsychotic medication prescription among adults with intellectual disabilities in the current study was much higher than in the general population, which is less than 1% (Marston et al., 2014). Whereas 35-37% of the adults with intellectual disabilities were prescribed antipsychotics in the current study, only 7.4-7.8% had a diagnosis of psychosis and/or bipolar disorder (severe mental illness) for which antipsychotics are licensed. This shows that in the vast majority of cases, antipsychotics were not prescribed for severe mental illness. The prevalence of antidepressant prescribing in the current study was much higher than the general population, which is 10.3% (NHS Digital, 2021). Whereas 17-25% of the adults with intellectual disabilities were prescribed antidepressants, only 3% had a diagnosis of depressive disorder in the current study for which antidepressants are licensed. This shows that in the vast majority of cases, antidepressants were not prescribed for depressive disorder.

### The incidence of new psychotropic prescriptions among adults with intellectual disabilities between 2015 and 2022

The incidence of new prescriptions of any psychotropic medications was 503 in 1999 and 533 per 10,000 person-years in 2013 in the English study (Sheehan et al., 2015), compared with the incidence rate of new psychotropic medications prescriptions of 457 in 2015 and 383 per 10,000 person-years in 2022 in the current study, which was lower than the rates in the English study at both data entry points. Whereas the English study (Sheehan et al., 2015) showed an increase in the incidence rate of new prescriptions for most psychotropic medications over 15 years, the current study showed a decrease in the incidence rate of most psychotropic medication prescriptions over seven years, except for an increase in hypnotic/sedative prescriptions. Despite this decrease in the incidence rate in the current study, apart from a reduction in anxiolytic prescriptions, the prevalence of antipsychotics, antidepressants, hypnotics/sedatives, and mood stabilisers increased significantly between 2010 and 2022. This is likely due to the long-term use of these medications. The reasons for this require urgent scrutiny to prevent inappropriate long-term prescribing of psychotropic medications in adults with intellectual disabilities.

However, Figure 2 shows a rise in incidences in almost all psychotropic medications around 2016, followed by a subsequent fall in 2017, and another rise in 2022, which is the last year of data collection in the current study. This increase is difficult to explain and may not directly relate to the COVID-19 effect (Gil-Llario et al., 2023). A similar trend was reported in the English study (Sheehan et al., 2015), in that after a reduction in 2010-11 in the incidence of new prescriptions of anxiolytics and antidepressants, there was a subsequent increase in 2012-13. However, it is difficult to understand why the incidence of new anxiolytic prescriptions decreased in the current study between 2015 and 2022 when the prevalence of anxiety disorder diagnosis significantly increased over the same period. One possible explanation is that more clinicians may be using SSRIs instead of anxiolytics to treat anxiety disorders in recent years.

### The prevalence of psychotropic medication prescribing between 2010 and 2022 among adults with intellectual disabilities who showed challenging behaviour

The prevalence of antipsychotic prescriptions among those who showed challenging behaviour in the current study was higher than that reported in the English study (Sheehan et al., 2015). For example, 47% of those with challenging behaviour received antipsychotics in the English study (Sheehan et al., 2015) compared with 62% in 2010 and 63% in 2022 in the current study. One possible explanation is that there is a relative lack of availability of non-pharmacological interventions for challenging behaviour in the Czech Republic compared to the UK.

In previous studies, among those who received psychotropic medications, most received them outside their licensed indications (Holden and Gitlesen, 2004; de Kuijper et al., 2010; Sheehan et al., 2015; Deb, 2024; Deb et al., 2025). For example, in the current study, antipsychotics were prescribed for severe mental illness only in 18% of cases in 2010 and 19% in 2022, compared with 24% in the Norwegian (Holden and Gitlesen, 2004), 22% in the Dutch (de Kuijper et al., 2010), and 29% in the English study (Sheehan et al., 2015). A similar trend has recently been reported among adults with intellectual disabilities in community settings in England and Wales, where antipsychotics were prescribed for severe mental illness only in 19% of cases (Deb et al., 2025). This means in over 80% of cases when antipsychotics were prescribed off-label for adults with intellectual disabilities, there was no record of any severe mental illness for which antipsychotics are indicated. However, among those who received antipsychotic medications, the proportion for whom they were used for challenging behaviour in the current study was lower (51-52%) than the Dutch study (58%) (de Kuijper et al., 2010) and the English study (61%) (Sheehan et al., 2015). Like the previous studies (Holden and Gitlesen, 2004; de Kuijper et al., 2010; Sheehan et al., 2015), the current study clearly showed that antipsychotic medications were prescribed primarily off-license for challenging behaviour in adults with intellectual disabilities rather than mental illnesses for which they are licensed.

A similar trend was observed for other psychotropic medications in the current study. For example, antidepressants were prescribed for depressive disorder only in 8-11% of cases but were prescribed more often for challenging behaviour (33-40% of cases). Similarly, mood stabilisers were prescribed for bipolar disorder only in 1% of cases, whereas they were prescribed for challenging behaviour in 38-40% of cases (see Table 2). This compares with a recent community-based study of adults with intellectual disabilities, which showed antiepileptic medications were used for epilepsy less often (in 32% of cases) than challenging behaviour (in 40% of cases) (Deb et al., 2025). This shows that psychotropic medications are prescribed off-licence in a high proportion of cases, particularly for challenging behaviour in adults with intellectual disabilities against the NICE and the WPA guidelines recommendations (Deb et al., 2009; NICE, 2015; Deb, 2024). However, the off-licence use of a licenced medication is not necessarily inappropriate if appropriate safeguards are implemented (Aitchison et al., 2013).

### The incidence of new psychotropic medication prescriptions among adults with intellectual disabilities between 2015 and 2022 who showed challenging behaviours

However, the incidence of new prescriptions for challenging behaviour showed, despite a decrease in overall psychotropic prescribing along with antipsychotics, antidepressants, and anxiolytics, an increase in the hypnotics/sedatives and mood stabiliser prescriptions between 2015 and 2022. This shows that in the current study, the welcome decrease in new prescriptions for antipsychotics, antidepressants, and anxiolytics for treating challenging behaviour may have been achieved at the expense of a not so welcome rise in other psychotropic prescriptions, hypnotics/sedatives and mood stabilisers. An English paper from the UK reported a recent trend in increased antidepressant prescribing to compensate for the reduction in antipsychotic prescribing (Branford & Shankar, 2022).

### Relationship among mental illnesses, challenging behaviour and psychotropic prescribing

The findings of a significant association among challenging behaviour and psychoses, bipolar, anxiety and personality disorders (see Table 4) are similar to what has been reported previously (Deb et al., 2022a, b). The incidences of psychoses, anxiety and personality disorders showed a significant association with more severe intellectual disabilities. However, diagnosis of mental illnesses, particularly schizophrenia, is unreliable in adults with severe and profound intellectual disabilities (see Deb et al., 2022a for a review). Similarly, challenging behaviour may have been diagnosed as anxiety and personality disorders in some cases, particularly among those with mild to moderate intellectual disabilities (Deb et al., 2022a, b).

### The relationship between psychotropic medication prescribing and other variables

Table 5 showed that the incidences of psychotropic medication prescriptions were significantly higher in the presence of mental illness, challenging behaviour, older age and more severe intellectual disabilities. A higher incidence of psychotropic prescribing in those with a mental illness and challenging behaviour had been reported before (de Kuijper et al., 2010; Sheehan et al., 2015; Deb, 2024). The significant association in the incidence of psychotropic medication prescribing and adults with severe intellectual disabilities and older age could have been a confounding effect of a similar association of age and severity of intellectual disabilities with mental illnesses and challenging behaviour. The fact that the female gender in the current study was statistically significantly associated with the prescription of antidepressants and anxiolytics but not antipsychotics and mood stabilisers is difficult to explain. It is possible that clinicians consider anxiety and depression as underlying causes of challenging behaviour, more so among women with intellectual disabilities than men. A statistically significant association between older adults with intellectual disabilities and psychotropic medication prescribing is in keeping with previous studies among older adults with intellectual disabilities (Odalović et al., 2024).

## Strengths and limitations

This is the first-ever population-based cohort study using a large national health database and ICD-10 of incidence and prevalence of mental illness, challenging behaviour and psychotropic medication prescribing and their relationship in adults with intellectual disabilities. No other large national health records-based study has used ICD criteria to diagnose mental illness and challenging behaviour before. Only one similar study in England used Reed codes from a general practice database to diagnose mental illness and challenging behaviour, which did not correspond with ICD-10 codes. They also used the British National Formulary rather than the WHO ATC for psychotropic medication classification used in this study. Therefore, it is difficult to generalise their findings outside England. No other large-scale study exists outside England, particularly in Eastern Europe. This is also the first study to assess the impact of WPA international, NICE guidelines, and the STOMP initiative in England, UK, on psychotropic medication prescribing for adults with intellectual disabilities.

Capturing data from the whole adult population with intellectual disabilities will never be possible as many adults with mild intellectual disabilities are not known to the services or do not receive the diagnosis of intellectual disabilities. In the current study, we aimed to be as inclusive as possible by capturing both primary and secondary diagnoses from psychiatric, medical, surgical, and other inpatient and outpatient settings, as well as laboratory and allied professional data. The total Czech adult population (18 years and older) was 8,688,840 in 2022 (data gathered from the Institute of Health Information and Statistics of the Czech Republic, provided by authors JJ and HM), and we captured data on 62,636 adults with intellectual disabilities. This gives a prevalence of 0·7% of intellectual disabilities in the Czech adult population. The prevalence of adults with intellectual disabilities is reported to be between 0·5% and 0·7% of the whole adult general population (Wittchen et al, 2011; Carey et al., 2016; Maulik et al., 2022; NHS Digital, 2021). The demographics of the intellectual disabilities’ population in 2022 in the current study showed a higher proportion (54%) of males, younger age, and mild to moderate intellectual disabilities, all of which are in keeping with what would be expected from an intellectual disabilities’ population-based sample (Carey et al., 2016; Maulik et al., 2022). Considering all these factors, we believe our sample is as representative of the total adult Czech population with intellectual disabilities as possible.

## Conclusion

Despite the reduction in incidences of mental illness and challenging behaviour, the significant rise in their prevalence over 12 years may indicate the chronic nature of these conditions. The rate of challenging behaviour significantly correlated with mental illnesses. The rate of psychotropic medication prescribing was significantly correlated with the rate of mental illness and challenging behaviour. There is a high prevalence of psychotropic medication use among adults with intellectual disabilities in the Czech Republic. The prevalence of most psychotropic medication prescriptions increased statistically significantly between 2010 and 2022, despite a decrease in the incidence rates of new prescriptions of most psychotropic medications, apart from hypnotics/sedatives, which increased between 2015 and 2022. Most psychotropic medications were prescribed only in a minority of cases for mental illnesses for which they are licensed. In a high proportion of cases, psychotropic medications were prescribed off-licence for challenging behaviour. Despite a reduction in new prescriptions of antipsychotics, antidepressants, and anxiolytics between 2015 and 2022 for adults with intellectual disabilities who showed challenging behaviour, which may be an impact of WPA and NICE guidelines, there was an increased rate of new prescriptions for hypnotics/sedatives and mood stabilisers. It, therefore, appears that the decrease in new prescriptions of antipsychotics, antidepressants, and anxiolytics may have been achieved at the expense of increased new prescriptions of hypnotics/sedatives and mood stabilisers. This is a worrying trend which requires further investigation. Similarly, the reasons for the significant rise in the prevalence rates of most psychotropic medications between 2010 and 2022, which goes against WPA and NICE guidelines, require stringent scrutiny to prevent future inappropriate overmedication of adults with intellectual disabilities, which is a significant public health concern.

## Contributors

Concept and design: Deb, Třešňák, Holub; Data acquisition, analysis, or interpretation of data: Jarkovský, Melicharová, Limbu, Deb; Drafting of the manuscript: Deb; Critical review of the manuscript: Deb, Jarkovský, Melicharová, Holub, Limbu, Třešňák. All authors approved the final version of the submitted manuscript.

## Funding

This study was not supported by any external funding.

## Ethical approval

Data in the National Health Information System (NHIS) are collected in compliance with Act No. 372/2011 Coll., on Health Services and Conditions of Their Provision. Due to this legal mandate, the retrospective analyses of data in our study did not require either an ethics committee approval or participants’ informed consent.

## Data Availability

Data are available from the Institute of Health Information and Statistics of the Czech Republic subject to appropriate authorization

## Notes

### Competing Interest Statement

The authors have declared no competing interest.

### Funding Statement

This study did not receive any funding

### Author Declarations

Institute of Health Information and Statistics of the Czech Republic. Unidentified group-level data were analysed for this study.

### Summary of Updates

We have added a new citation for Holden and Gitlesen (2004) and changed a figure concerning the reference of Sheehan et al. (2015).

